# The effectiveness of a complex intervention, aimed at reducing hospital occupancy, to improve Emergency Department patient flow: a retrospective controlled interrupted time series

**DOI:** 10.64898/2026.08.31.26361802

**Authors:** Ryan McHenry, David Caesar, Benjamin Clarke, Daniel Mackay, Jill Pell

## Abstract

**Objectives:** Emergency department (ED) crowding is recognised as an important public health concern internationally, and is driven principally by exit block, the shortage of inpatient beds for patients requiring admission. This study aimed to evaluate whether a complex intervention targeting hospital occupancy improved ED patient flow, and quantified the change in attendances.

**Methods:** A controlled interrupted time series using weekly, publicly reported Public Health Scotland data from 1 January 2022 to 1 February 2026. The multi-component intervention focused on reducing hospital occupancy and included additional adult social care funding; engagement with regional social care providers; accelerated implementation of the Discharge without Delay programme; re-evaluation of whole-hospital escalation thresholds and response; resource and data supporting inpatient department reductions in length of stay; and additional investment in remote clinical assessment. The intervention commenced at a large tertiary ED on 01 February 2025. Primary outcomes were the proportions of attendances spending ≥4, ≥8 and ≥12 hours in the ED. The secondary outcome was attendance volume. Segmented regression was fitted with a contemporaneous control series, seasonal terms and autoregressive moving average errors. Long waits were additionally illustrated as potentially avoided deaths.

**Results:** The analysis covered 161 pre-intervention and 52 post-intervention weeks. Relative to pre-intervention levels, the proportion of attendances waiting over 4 hours fell by 10.4% (95% CI 1.6 to 19.2%), by 16.4% (95%CI 1.3 to 31.5%) over 8 hours and by 24.3% (95%CI 2.6 to 46.1%) over 12 hours. Using established associations between long ED waits and excess mortality, by one-year the intervention was potentially associated with 54 fewer excess deaths (95%CI 19 to 93). Attendances rose by 3.8% (95%CI 1.3 to 6.4%) against the counterfactual.

**Conclusions:** A complex intervention targeting hospital occupancy was associated with a reduction in long ED waits despite rising attendances. Interventions addressing hospital occupancy can meaningfully improve ED crowding.

---

Emergency Department (ED) crowding is an increasing international public health concern, and has been associated with patients harms, including avoidable deaths.[1,2] Crowding has mainly been attributed to output issues such as ‘exit block’, poor access to inpatient beds for those requiring admission.[2–4] This itself is often caused by delayed discharge for inpatients, longer inpatient length of stay and higher hospital occupancy.[5,6]

While some healthcare systems and interventions have aimed to mitigate the harms of crowding through measures that target throughput (those aiming to reduce patient processing times within the ED), or input (those that seek to reduce presentations to emergency care), [3] in the context of constraints related to access to inpatient beds, it is likely that only those that target the root cause can sustainably improve crowding.[7] Prior evidence suggests that interventions focussed on improving the timeliness of discharge from hospital have the potential improve ED crowding.[6,8] However, even in systems with wide implementation of such interventions, [9,10] crowding remains common and associated with significant numbers of excess deaths.[1,2,11]

As part of efforts to improve the performance of unscheduled care in the National Health Service (NHS) Lothian region, the Scottish Government invested £14.5 million [12], focussed on the Royal Infirmary of Edinburgh site. This investment was used to implement changes designed to reduce whole-system occupancy, predominantly through funding for adult social care placements, admission avoidance strategies through an ED frailty service, and a whole-system commitment to reducing inpatient length of stay.

As ED crowding is an international concern, understanding which interventions succeed, and by what magnitude, is an important public health priority.[11,13,14] This study aimed to assess the extent to which a complex intervention, aimed at reducing hospital occupancy, was associated with changes in ED patient flow, as assessed by ED performance metrics, the proportion of patients waiting ≥4, ≥8 and ≥12 hours for admission or discharge; it also aimed to assess whether the intervention changed the number of ED attendances. Furthermore, the study also aimed to demonstrate the feasibility of applying econometric methodology to routinely-reported public data to evaluate the impact of operational interventions in emergency and acute care.

## Methods

### Setting and intervention

The intervention was instituted in the Royal Infirmary of Edinburgh at Little France (RIE), Edinburgh, Scotland, United Kingdom in February 2025. The RIE is a major tertiary adult (age ≥16 years) hospital and Major Trauma Centre within the health board, NHS Lothian. The RIE ED has approximately 125,000 annual attendances and serves a core population of 920,000.[15]

The Lothian Strategic Development Framework (LSDF) aimed to improve the performance of unscheduled care by reducing ED attendances, reducing hospital occupancy and length of stay, and reducing inappropriate admissions. [12] The majority of funding was targeted at improving community capacity for social care. In-hospital, the intervention consisted of accelerated implementation of the Discharge without Delay (DwD) programme, including rapid adoption of planned date of discharge (PDD);[16] hospital-wide re-evaluation of escalation thresholds and actions on meeting these thresholds; engagement across senior leadership and social care partners; and strengthening of an ED-based frailty service.[17] In Scotland, territorial health boards hosting hospitals interact with local authority partners that support social care (Health and Social Care Partnerships; HSCPs). Each HSCP in the intervention region was provided with targets for their own inpatient residents, aiming to reduce delayed discharges by the provision of expedited social care in conjunction with funding for social care itself. Inpatient departments were provided with resources to reduce length of stay, and department-level data provision on length of stay was improved, including length of stay metrics disaggregated by individual hospital ward. [18]

Funding was also provided to the Flow Navigation Centre, a virtual service offering clinical decision-making introduced as part of measures to reduce the number of patients self-presenting to EDs in Scotland.[19] This service is accessible to clinicians in community settings, such as paramedics and general practitioners, as well as members of the public who may be referred via the existing telephone advice service, NHS 24.[20] The interventions were instituted at the beginning of February 2025.

### Data and outcomes

Public Health Scotland publishes weekly, site-level data on ED performance, including the number of presentations, and the number of patients spending more than 4-, 8- and 12-hours in EDs before admission, discharge or transfer.[21] Data were considered from 1^st^ January 2022 (3 years prior to the intervention) to 1^st^ February 2026 (1 year following the intervention) for the host site and control EDs.

As there is evidence that longer ED length of stay is associated with excess mortality,[1] the primary outcomes were the proportion of patients waiting ≥4, ≥8 and ≥12 hours for discharge, admission, or transfer from the ED. As a stated strategic aim of the plan, the secondary outcome was changes in the number of attendances at the ED.[12]

Published thresholds are cumulative, therefore every patient waiting ≥12 hours also appears in the ≥8 and ≥4 hour counts. To describe where changes occurred, and to avoid counting the same patient more than once, we also derived three non-overlapping bands (4–8 hours, 8–12 hours, over 12 hours) by differencing the published counts, and analysed each for outcomes concerning avoided admissions and deaths.

Hospital length of stay is routinely reported on a quarterly basis in Scotland.[22] As the proposed mechanism of effect for this intervention, changes in mean length of stay between the intervention site, and the control sites, are therefore reported descriptively.

### Design and analyses

Controlled interrupted time series (CITS) design was used to assess the post-intervention step-change in the outcomes of interest. Interrupted time series is widely used to evaluate natural experiments, since randomisation is not feasible.[23] CITS design augments a conventional interrupted time series with a control series, so that the counterfactual, what the treated site would have experienced without the intervention, is anchored to contemporaneous changes shared across sites (for example, seasonal pressures and national policy interventions) rather than to within-site trends alone. This mitigates the principal threat to a single-series interrupted time series, where co-occurring system-wide change is mistaken for an intervention effect.[24,25]

Control sites were selected using pre-intervention data only, so that selection could not be influenced by the post-intervention outcome. Candidate controls were ranked by the correlation of their pre-period residuals with those of the treated site, and the number of controls was chosen to minimise the Bayesian information criterion of the treated-site pre-period model.

The intervention effect was estimated as a level change in the outcome series at the intervention commencement date. The segmented-regression was a difference-series where the difference between the treated-site outcome and the mean of the controls was regressed on trend and seasonality terms, and a binary intervention variable.

Seasonality was represented by two Fourier harmonic pairs at a period of 52.18 weeks. Serial correlation in the residuals was modelled by an autoregressive moving average (ARMA 1,1) error structure, and models were fitted by restricted maximum likelihood using generalised least squares. Effect estimates are reported as percentage-point changes with 95% confidence intervals (95%CIs).

Because a complex intervention rarely achieves full effect at initiation,[25] transition sensitivity analyses were used; a washout excluding the first 4, 8 or 13 weeks, and a linear ramp reaching full effect over the same periods. Robustness was further assessed against a simpler autoregressive (AR1) error structure and exclusion of control sites sharing a health board with the treated site (to guard against contamination or diversion).

A Bayesian structural time series model on the weekly counts for each waiting time band and attendances were used as a further sensitivity analysis, using the same control pool, conservative local-level trends and weakly informative spike-and-slab priors.[26] Structural break assumptions were examined using an unsupervised Bayesian changepoint estimator, and residuals assessed by Ljung–Box test and autocorrelation plots. Full specification, sensitivity analyses and diagnostics are provided in the online supplement.

### Avoided long-wait admissions and excess deaths

There is an established association between long ED waits amongst patients admitted to hospital and harms, though less evidence on those who are discharged following emergency attendance.[1] The number of long wait *admissions* was determined by adjusting the number waiting at each breach band by a fixed ratio, determined as the proportion of patients admitted in each band in a Scottish dataset, this was 40%, 70% and 80% admissions at the 4-8, 8-12, and ≥12-hour band respectively. The number of long wait admissions potentially avoided was calculated from the observed-minus-counterfactual gap for each waiting time band.

Using the established association between long ED waits and excess deaths in admitted patients determined by Jones et al.[1], avoided deaths were estimated by Monte Carlo propagation, drawing avoided admissions from their sampling distribution and the number needed to harm from a log-normal fitted to the published confidence interval. As routine data do not disaggregate 6–8 hour waits, all 4–8 hour waits were conservatively assigned the 4–6 hour magnitude of harm; and as reliable associations with mortality for those waiting ≥12 hours are not available, these waits were assigned the 8-hour figure. The same propagation was applied to the Bayesian structural time series sensitivity analysis.

### Statistical power

The study was simulated using Public Health Scotland data with 2000 bootstrap replications. At a power of 0.8 and alpha of 0.05, the minimum detectable effect for absolute change in the percentage of patients spending more than 4 hours in the ED at the intervention site was 6.7%.[27]

### Analysis environment

As an evaluation of an already-instituted intervention using publicly-available data, the study was defined as service evaluation and did not require ethical review.[28] Statistical significance was set at p<0.05. Analyses were conducted using the nlme, Rbeast and CausalImpact packages in in R (Version 4.5.0).[26,29,30]

## Results

From 1^st^ January 2022 to 1^st^ February 2026, there were 505,921 presentations to the RIE ED, summary statistics of these presentations are demonstrated in Table 1.

**Table 1.**
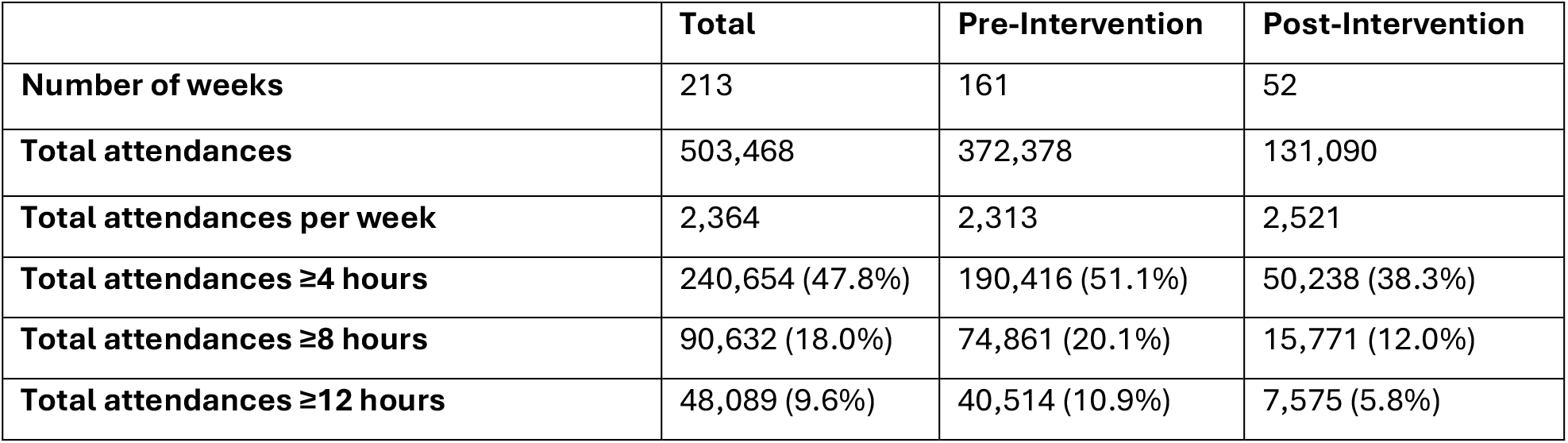
Summary statistics for the number of presentations to the intervention site before and after the intervention, including stratification by 4-, 8- and 12-hour wait status.

Mean hospital length of stay fell from 11.8 days in the quarter in which the intervention was introduced, to 9.6 days in the following quarter (Supplementary Figure 1).

Following the intervention the proportion of attendances waiting more than 4 hours fell by 5.3 percentage points (95%CI 0.8 to 9.8) relative to the control-based counterfactual. Reductions were also seen at 8 hours (3.3 percentage points; 95%CI 0.3 to 6.4) and 12 hours (2.7 percentage points; 95% CI 0.3 to 5.0). Absolute effect estimates for all thresholds and bands are given in Table 2 alongside the relative change in comparison to pre-intervention rates, and demonstrated in Figure 1.

**Figure 1.**
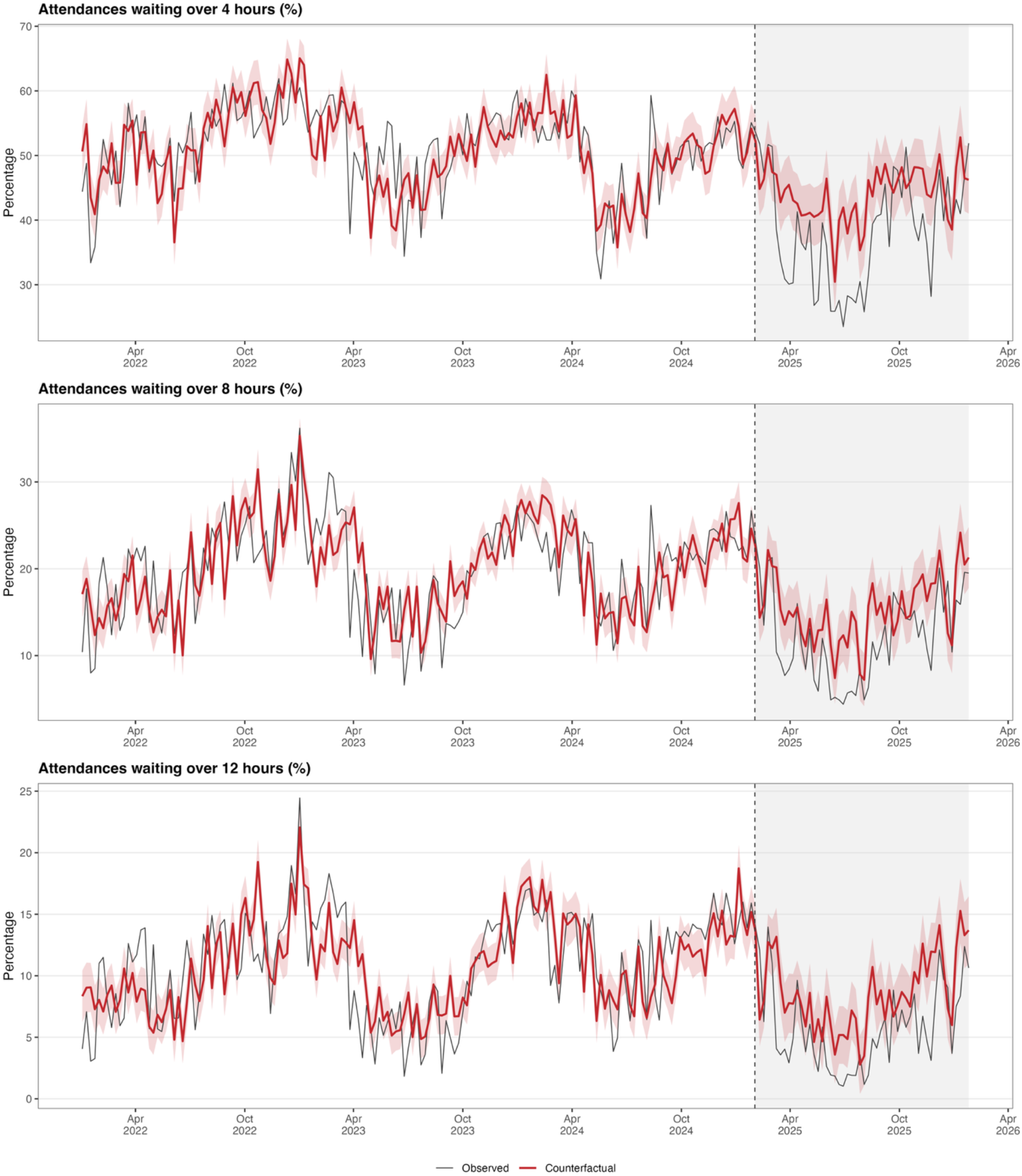
Proportion of Emergency Department attendees waiting ≥4, ≥8, and ≥12 hours, observed and counterfactual.

**Table 2.** Step change in the proportion of attendances breaching each waiting time threshold and band. The pre-intervention rates do not exactly sum due to rounding.

| Outcome | Pre-intervention rate | Absolute change (95% CI) | Relative change (95% CI) |
| --- | --- | --- | --- |
| **Waiting time thresholds** |  |  |  |
| ≥4 hours | 51.1% | -5.32 (95% CI -9.83 to -0.82) | -10.4% (95% CI -19.2 to -1.6) |
| ≥8 hours | 20.1% | -3.31 (95% CI -6.36 to -0.27) | -16.4% (95% CI -31.5 to -1.3) |
| ≥12 hours | 10.9% | -2.66 (95% CI -5.04 to -0.28) | -24.3% (95% CI -46.1 to -2.6) |
| **Waiting time bands** |  |  |  |
| 4 to 8 hours | 31.1% | -1.12 (95% CI -5.04 to 2.80) | -3.6% (95% CI -16.2 to 9.0) |
| 8 to 12 hours | 9.2% | -0.58 (95% CI -1.96 to 0.80) | -6.2% (95% CI -21.2 to 8.7) |
| ≥12 hours | 10.9% | -2.66 (95% CI -5.04 to -0.28) | -24.3% (95% CI -46.1 to -2.6) |

Analysis by non-overlapping waiting time bands showed the improvement was not evenly distributed (Figure 2): the absolute change for the 4 to 8 hour band was -1.1 percentage points (95% CI -5.0 to +2.8); for 8 to 12 hours, -0.6 percentage points (95% CI -2.0 to +0.8); and for those over 12 hours, -2.7 percentage points (95% CI -5.0 to - 0.3). The absolute and relative change is shown in Table 2 and the relative change for each band is visualised in Figure 2.

**Figure 2.**
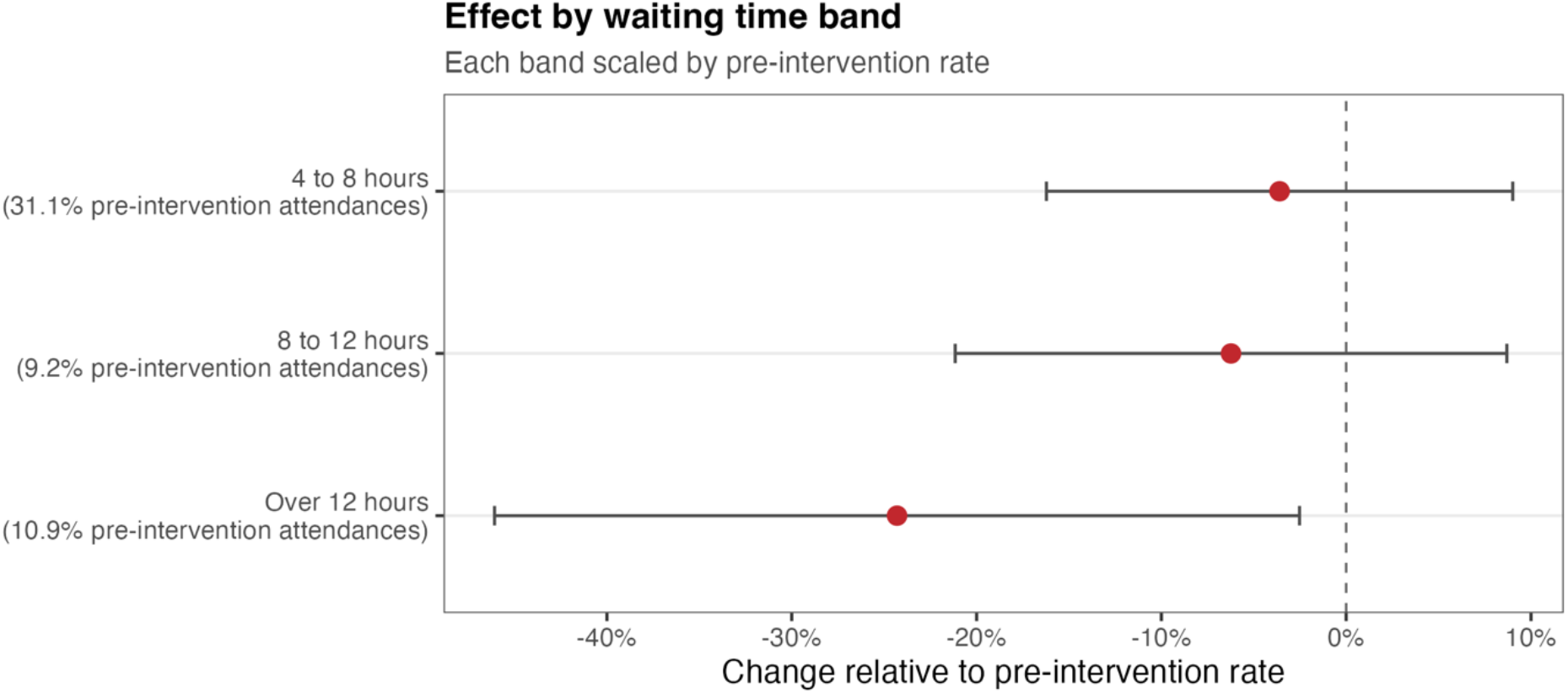
Relative change in each non-overlapping wait band, expressed relative to that band’s pre-intervention rate, with 95% confidence intervals.

Applied to the gap between observed and expected attendance volumes, there were 7,149 (95%CI 1,718 to 12, 580) fewer attendees waiting ≥4 hours across the year following the intervention, summed across the three bands, of which it is estimated 4,561 (95%CI 1,607 to 7,516) would have been patients awaiting an inpatient bed (Table 3).

**Table 3.** Long waits and long-wait admissions avoided over year following the introduction of the intervention, by wait bands. Calculated by the observed-minus-counterfactual gap for each threshold.

| Wait band | Long waits averted (95% CI) | Avoided long-wait admissions (95% CI) |
| --- | --- | --- |
| 4 to 8 hours | 2,699 (95% CI -1,822 to 7,219) | 1,079 (95% CI -729 to 2,888) |
| 8 to 12 hours | 787 (95% CI -715 to 2,288) | 551 (95% CI -501 to 1,602) |
| Over 12 hours | 3,664 (95% CI 1,055 to 6,272) | 2,931 (95% CI 844 to 5,018) |
| All waits over 4 hours (sum of bands) | 7,149 (95% CI 1,718 to 12,580) | 4,561 (95% CI 1,607 to 7,516) |

There was no significant change in slope for any outcome. Cumulative averted delays are demonstrated in Supplementary Figure 2, and slope changes are described in Supplementary Table 1 and Supplementary Figure 3.

Applying the association reported by Jones *et al.[1]* to the observed shortfall in long-wait admissions gives an estimated 54 (95%CI 19 to 93) fewer deaths over the year.

ED attendances rose by 3.8% (95% CI 1.3 to 6.4%) relative to the counterfactual (Figure 3).

**Figure 3.**
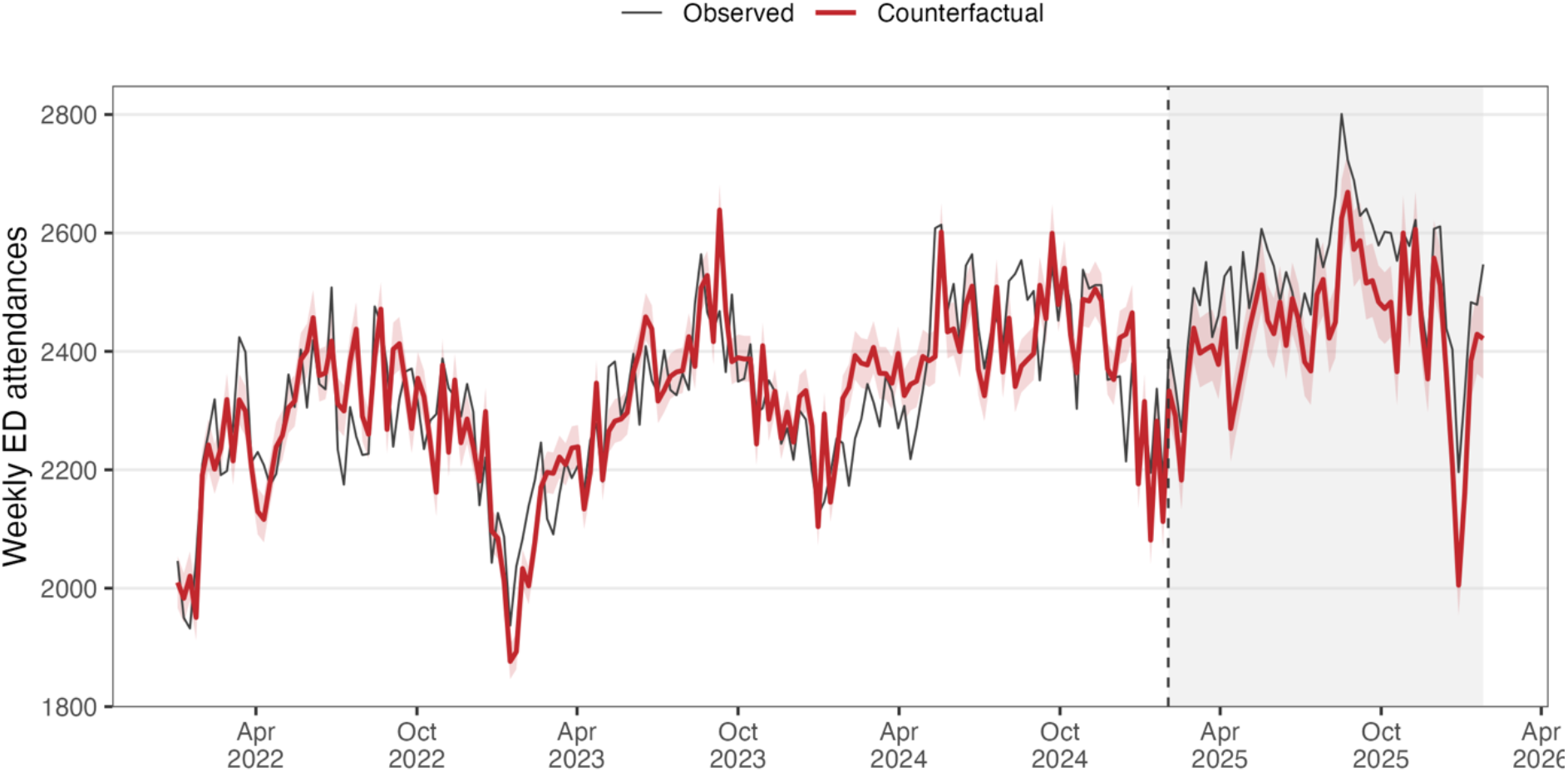
Weekly ED attendances, observed and counterfactual.

Bayesian structural time series analyses confirmed the direction of effect of the primary analyses; although the 95% credible interval excluded the null for each of the analyses and demonstrated a greater magnitude of effect for each of the outcomes. Using this modelling, the number of excess deaths avoided during the year following the intervention was estimated as 103 (95% Credible interval 78 to 133) (Supplementary Tables 2 & 3 and Supplementary Figure 4A-D).

Bayesian changepoint analysis demonstrated a structural break shortly following the intervention (Supplementary Figure 5). The primary estimates were robust to sensitivity analyses allowing 4, 8 or 13 weeks for the effect to establish, by washout or linear ramp. Full model outputs, and sensitivity analyses are demonstrated in Supplementary Tables 4 & 5. Ljung-Box tests demonstrated no significant residual autocorrelation (p>0.05).

ACF and PACF plots demonstrated no significant residual autocorrelation for the waiting time thresholds (Supplementary Figure 6).

## Discussion

This study evaluating the effectiveness of a complex intervention, aiming to improve waiting times in emergency care through reductions in inpatient length of stay, demonstrated significant improvements in ED performance as measured by the proportion of patients with an ED length-of-stay of ≥4, ≥8 and ≥12 hours. Applying the current best estimates of harm from long ED waits, it is likely that this intervention also reduced avoidable deaths from long waits in emergency care. These benefits were achieved despite an increase in ED attendances.

Analysis by waiting time band demonstrates a pattern of change that such interventions might be expected to exert on long ED waits. The benefit was concentrated among the longest waits, with little net change in the 4 to 8 hour, or 8 to 12 hour wait band. This is intuitive for an intervention acting on exit block: improving ED access to inpatient beds shortens the extreme tail of ED delays, moving patients out of the longest bands and into shorter ones. It also implies that reporting the 4-hour standard alone understates what such interventions achieve, because the standard is insensitive to redistribution within the breaching population, and undervalues a reduction in longer waits where the mortality burden is concentrated.

Despite strategic aims, and funding for efforts, to reduce ED presentations, these increased in the post-intervention period. While it is reassuring that such increases did not result in poorer ED performance, it also reflects the mechanistic reality of contemporary ED crowding, which results from ‘output’ issues rather than ‘input’ or demand. [3,13] It is possible that some of the intervention mechanisms in fact increased ED presentations. The staffing of remote clinical advice centres may represent supply-sensitive care, where increased supply results in increased resource use, even where this does not bring value to the patient or healthcare system.[31]

Emergency Departments in the United Kingdom and internationally are under severe system strain. These findings, of the real-world effectiveness of a complex intervention targeting a system constraint, have the potential to inform healthcare systems to build sustainable and meaningful change. Further research is needed; to establish which components of the intervention are most effective, if they are cost-effective, if they are safe and acceptable for patients, and if they can be implemented in other settings.

### Strengths and Limitations

Interrupted time series is regarded as amongst the strongest quasi-experimental designs in settings where randomisation is not feasible, such as the natural experiment in this study.[23] The results demonstrate that such econometric analyses are feasible, can produce meaningful results, and when implemented carefully, can produce clinically- and operationally-significant minimum detectable effects, utilising publicly-available data.

The number of excess deaths potentially avoided due to the intervention are likely to be an underestimate as routinely-reported data does not disaggregate those waiting 6-8

hours who are known to be at greater risk than those waiting 4-6 hours, and no known estimates for those waiting ≥12 hours despite a linear association between long waits and mortality.[1] The analysis also does not account for potentially avoidable harm to those who experience long waits prior to being discharged from the ED, as previous studies exploring the association of mortality with overcrowding have been limited to admitted patients. Regardless, the conservative approach used in this study has precedent in reporting of the harms associated with ED crowding.[11] Jones et al. presented excess deaths as associations rather than causal findings, as such, the potential reductions in excess deaths framed in this work should be interpreted as representative modelled estimates of the effectiveness of the intervention rather than a measured mortality effect.[1]

Several limitations apply. This is a quasi-experimental design and cannot exclude an unmeasured change coinciding with implementation that affected the intervention site but not controls. The intervention was a bundle, so the analysis cannot identify which components were effective. Estimates are site-level; if improvement arose partly from diverting patients elsewhere, the site-level benefit would overstate system-wide gain, and the exclusion of same-board controls in sensitivity analyses addresses this only partially. Routine data carry no patient-level detail, so case mix and acuity are unobserved.

## Conclusion

This study of a complex intervention aimed at reducing hospital occupancy demonstrated reductions in ED crowding after its implementation. This occurred despite a concurrent increase in emergency presentations, suggesting that hospital occupancy, not presentations, is a major driver of ED crowding. The study shows that interventions targeting this systemic cause have the potential to meaningfully improve the experience of, and outcomes from, emergency care.

Future research could use similar methodologies to efficiently evaluate the real-world effectiveness of other interventions designed to mitigate the harms of ED crowding and long length of stay. Policymakers should recognise that the major driver of crowding is exit block, and ensure resource and interventions are directed to address it.

## Data Availability

All data is publicly available, hosted by Public Health Scotland

