## Supplement for "The effectiveness of a complex intervention, aimed at reducing hospital occupancy, to improve Emergency Department patient flow: a retrospective controlled interrupted time series"

**Supplementary Methods**

*Control selection*

Controls were selected on pre-intervention data only, so that selection could not be influenced by post-intervention outcomes. For each candidate site, the residuals of a regression of its pre-period series on time and Fourier terms were correlated with the equivalent residuals for the treated site. Candidates were ranked by that correlation, and the number retained chosen to minimise the Bayesian information criterion of the treated-site pre-period model.

The procedure selected: Forth Valley Royal Hospital, St John's Hospital. The same panel was applied to every outcome series, including the bands, so that band estimates are mutually comparable and comparable with the cumulative series. St John's Hospital is a same-board site, and so was excluded in sensitivity analyses (see below).

*Sensitivity analyses*

Two structural sensitivities were used: a simpler AR(1) error structure; and an exclusion of control sites sharing a health board with the treated site, to guard against contamination or diversion violating the stable-unit-treatment-value assumption.

Transition sensitivities were used to address phase-in issues. A complex intervention rarely achieves full effect at initiation; if the true effect phases in but the model imposes a sharp step, the step coefficient averages the transition with the steady state and understates the effect. Two transition forms were fitted for $w = 4$, 8 and 13 weeks: a washout, dropping the first $w$ post-intervention weeks from the likelihood entirely, which is robust to any transition shape at the cost of discarding information; and a linear ramp rising from zero at go-live to one at week $w$, which uses all data but assumes the phase-in is linear. Washout standard errors necessarily grow with $w$, as fewer post-intervention weeks contribute; a wider interval at $w = 13$ should not be read as a weaker effect.

*Attendance volume*

Attendance volume was modelled on the log scale, so the intervention coefficient is interpretable as a proportional change.

Sites sharing a health board with the treated site were excluded from the control pool for this outcome, because they are the sites to which patients would most plausibly be diverted. The control panel for attendances therefore differs from that used for the breach outcomes.

*Cumulative effects and attenuation*

The primary analysis was the effect as a level change. To characterise the effect across the follow-up year, including any attenuation, the weekly gap between the observed series and the model-based counterfactual was formed. A cumulative curve that rises then flattens indicates a waning effect, while a linear accumulation would indicate a sustained effect. A formal test regresses the weekly gap on time since implementation, with AR(1) errors.

*Structural break diagnostics*

A Bayesian estimator of abrupt change, seasonality and trend (BEAST) was applied to the treated and control series.[1] BEAST infers the number and location of trend changepoints, seasonal changepoints and outliers by Bayesian model averaging, returning a per-week posterior probability of a changepoint. It is unsupervised, and estimates no treatment effect. It was used to detect pre-period breaks that would impair counterfactual stability, breaks coincident with implementation in control sites, and to corroborate the break at the treated site.

*Residual diagnostics*

Residual autocorrelation was assessed by the Ljung–Box statistic on normalised residuals.

*Conversion to admissions and deaths*

Averted breaches were computed by applying the per-week estimated effect to observed attendance volumes across the follow-up window. The manuscript reports the observed-gap effect.

Avoided long-wait admissions were obtained by applying, to each non-overlapping band, the proportion of patients within that band admitted in Scottish data (40%, 70% and 80% at 4-8, 8-12 and over 12 hours respectively).

Avoided deaths were estimated by Monte Carlo propagation with 50,000 draws using the *Jones et al.* standardized mortality rate as discussed in the main manuscript.[2]


1 Zhao K, Wulder MA, Hu T, *et al.* Detecting change-point, trend, and seasonality in satellite time series data to track abrupt changes and nonlinear dynamics: A Bayesian ensemble algorithm. *Remote sensing of Environment*. 2019;232:111181.

2 Jones S, Moulton C, Swift S, *et al.* Association between delays to patient admission from the emergency department and all-cause 30-day mortality. *Emergency Medicine Journal*. 2022;39:168–73.

Supplementary Table 1. Slope changes for each of the wait threshold outcomes and attendances

| Outcome | Annual Slope Change | p value |
|---|---|---|
| Wait ≥4 hours | +1.66% (-8.01 to +11.33%) | 0.737 |
| Wait ≥8 hours | -0.43% (-7.10 to +6.24%) | 0.900 |
| Wait ≥12 hours | -2.85% (-7.96 to +2.26%) | 0.276 |
| Attendances | +2.12% (-3.22 to +7.76%) | 0.444 |

Supplementary Table 2. Bayesian structural time-series posterior estimates for the effect of the intervention on the weekly number in each waiting time band and attendances.

| Outcome | Average change (95%CrI) | Cumulative change (95%CrI) | Relative change % (95%CrI) | Bayesian posterior tail-area probability |
|---|---|---|---|---|
| 4 to 8 hours | -54.7 (-84.3 to -24.0) | -2847 (-4385 to -1250) | -7.6 (-11.3 to -3.5) | 0.005 |
| 8 to 12 hours | -60.3 (-76.4 to -44.1) | -3136 (-3975 to -2292) | -27.6 (-32.7 to -21.9) | <0.001 |
| Over 12 hours | -112.9 (-139.6 to -86.4) | -5871 (-7261 to -4492) | -43.5 (-48.9 to -37.2) | <0.001 |
| Attendances | 133.5 (95.4 to 169.6) | 6943 (4961 to 8818) | 5.6 (3.9 to 7.2) | <0.001 |

CrI – Credible Interval

Supplementary Table 3. Bayesian structural time-series estimates of averted long waits, avoided long-wait admissions and avoided deaths

| Wait band | Assumed admission fraction | Breaches averted (95% CrI) | Admissions avoided (95% CrI) | Deaths avoided (95% CrI) | Bayesian posterior tail-area probability |
|---|---|---|---|---|---|
| 4 to 8 hours | 0.4 | 2,849 (1,117 to 4,581) | 1,140 (447 to 1,833) | 5.9 (2.5 to 10.6) | 0.005 |
| 8 to 12 hours | 0.7 | 3,135 (2,291 to 3,979) | 2,195 (1,604 to 2,785) | 30.3 (20.0 to 43.9) | <0.001 |
| Over 12 hours | 0.8 | 5,876 (4,354 to 7,398) | 4,701 (3,483 to 5,919) | 64.9 (44.6 to 93.3) | <0.001 |
| Total | - | 11,860 (9,405 to 14,316) | 8,035 (6,515 to 9,556) | 101.9 (77.7 to 133.3) | <0.001 |

CrI – Credible Interval

Supplementary Table 4. Sensitivity analyses for wait times by threshold and attendances. Step change in percentage points (95% CI) at each cumulative threshold, by transition specification. 'Step' is the primary specification; 'washout' excludes the first w post-intervention weeks; 'ramp' phases the effect in linearly over w weeks.

| Specification | Over 4 hours | Over 8 hours | Over 12 hours | Attendances |
|---|---|---|---|---|
| Step | -5.32 (-9.83 to -0.82) | -3.31 (-6.36 to -0.27) | -2.66 (-5.04 to -0.28) | +3.79 (+1.26 to +6.38) |
| Washout, 4 weeks | -6.99 (-11.34 to -2.63) | -4.03 (-7.12 to -0.93) | -3.55 (-5.99 to -1.10) | +3.92 (+1.29 to +6.61) |
| Washout, 8 weeks | -7.49 (-12.01 to -2.97) | -3.99 (-7.19 to -0.78) | -3.40 (-5.94 to -0.86) | +4.05 (+1.31 to +6.87) |
| Washout, 13 weeks | -7.02 (-11.81 to -2.24) | -3.99 (-7.34 to -0.63) | -3.48 (-6.13 to -0.83) | +3.66 (+0.86 to +6.53) |
| Ramp, 4 weeks | -6.74 (-11.26 to -2.22) | -3.99 (-7.09 to -0.89) | -3.34 (-5.76 to -0.92) | +4.02 (+1.36 to +6.74) |
| Ramp, 8 weeks | -7.92 (-12.43 to -3.41) | -4.28 (-7.42 to -1.14) | -3.66 (-6.12 to -1.19) | +4.09 (+1.35 to +6.91) |
| Ramp, 13 weeks | -7.46 (-12.11 to -2.82) | -4.06 (-7.29 to -0.84) | -3.52 (-6.04 to -1.00) | +4.16 (+1.28 to +7.12) |

Supplementary Table 5. Structural sensitivity analyses for wait times by threshold and attendances. Step change in percentage points.

| Variant | Over 4 hours | Over 8 hours | Over 12 hours | Attendances |
|---|---|---|---|---|
| AR(1) errors | -5.94 (-9.64 to -2.24) | -3.46 (-6.09 to -0.84) | -2.89 (-4.87 to -0.92) | +3.98 (+1.74 to +6.27) |
| Same-board control excluded | -8.30 (-12.51 to -4.10) | -7.27 (-10.64 to -3.89) | -4.41 (-7.22 to -1.59) | +3.79 (+1.26 to +6.38) |

Supplementary Figure 1. Mean hospital length of stay by hospital site, including the intervention and control sites, in annual quarters. The grey shading indicates the quarter in which the intervention was initiated.

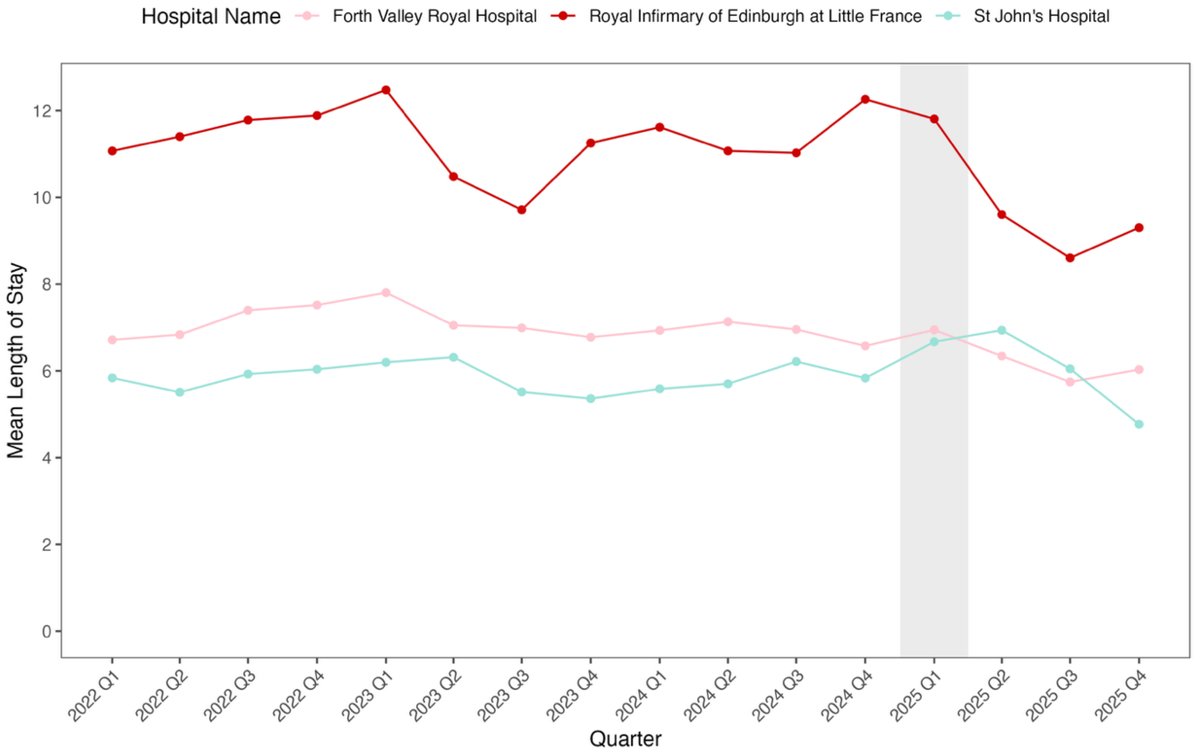


Supplementary Figure 2. Cumulative averted attendances by each wait-time threshold.

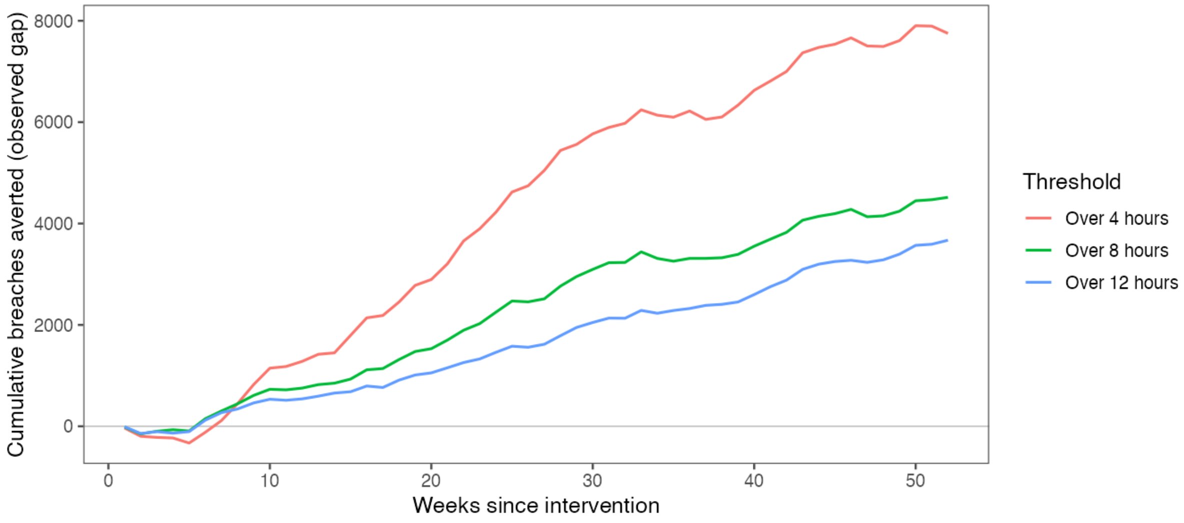

Supplementary Figure 3. Modelled effect at each week from implementation model with shaded 95% confidence band.

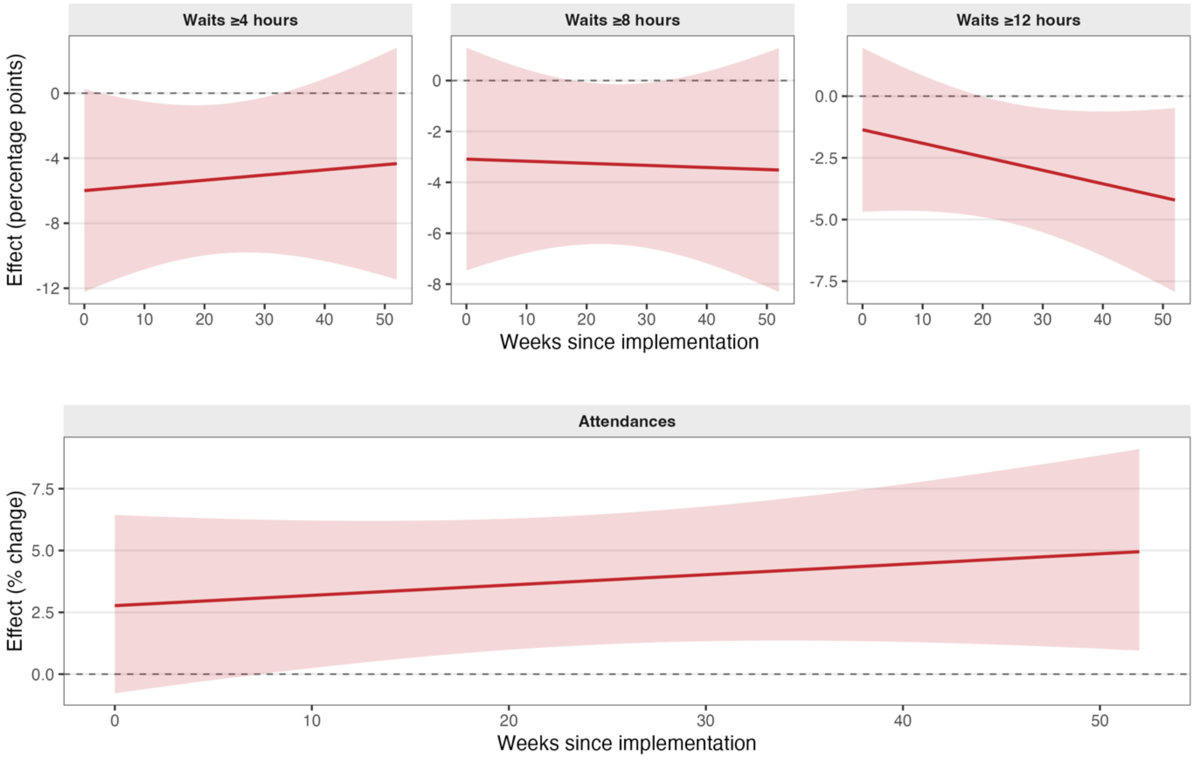

Supplementary Figure 4A-D. Visualisation of the Bayesian structural time series analysis of the effect of the intervention on the number of patients at each waiting time band (3A-C), and total number of attendances (3D). In the original time series, the black line is the observed value, the blue line is the counterfactual, and the shaded areas are the 95% Credible Interval. The pointwise and cumulative estimates give the site characteristics relative to the counterfactual (dashed line) and 95% Credible Intervals.

**4A**

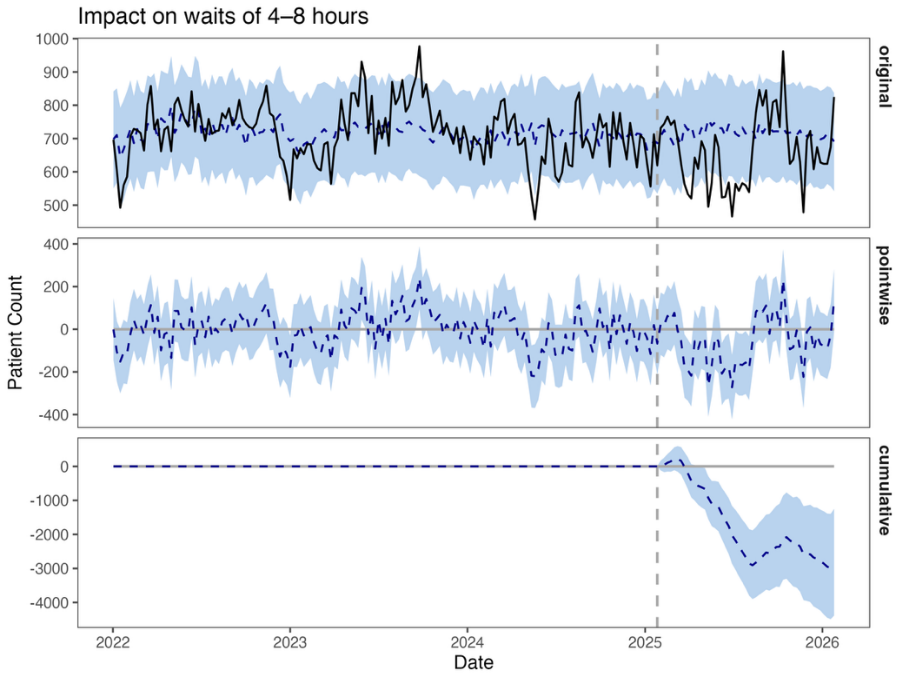


**4B**

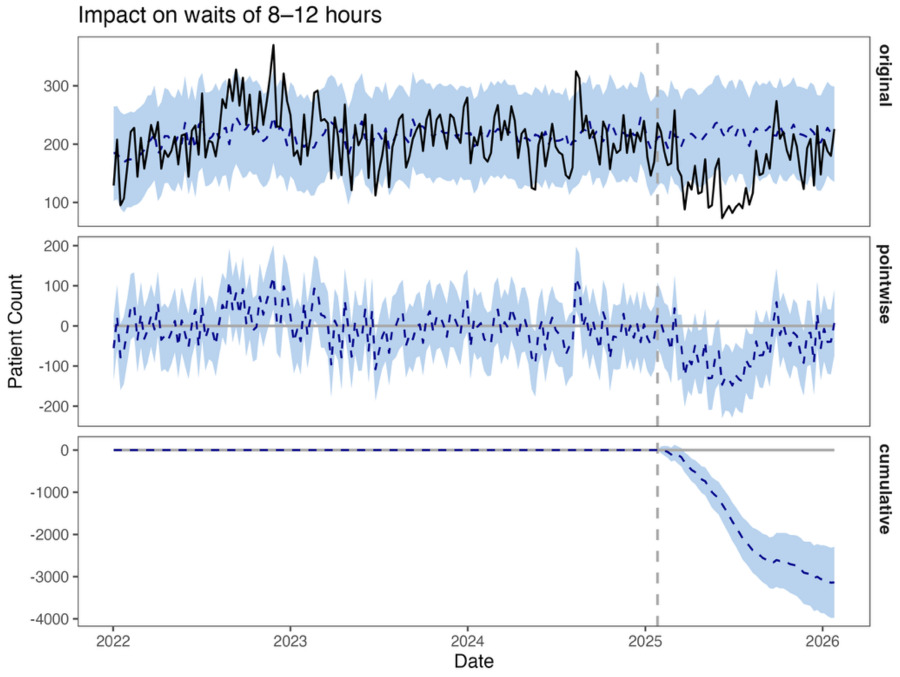

**4C**

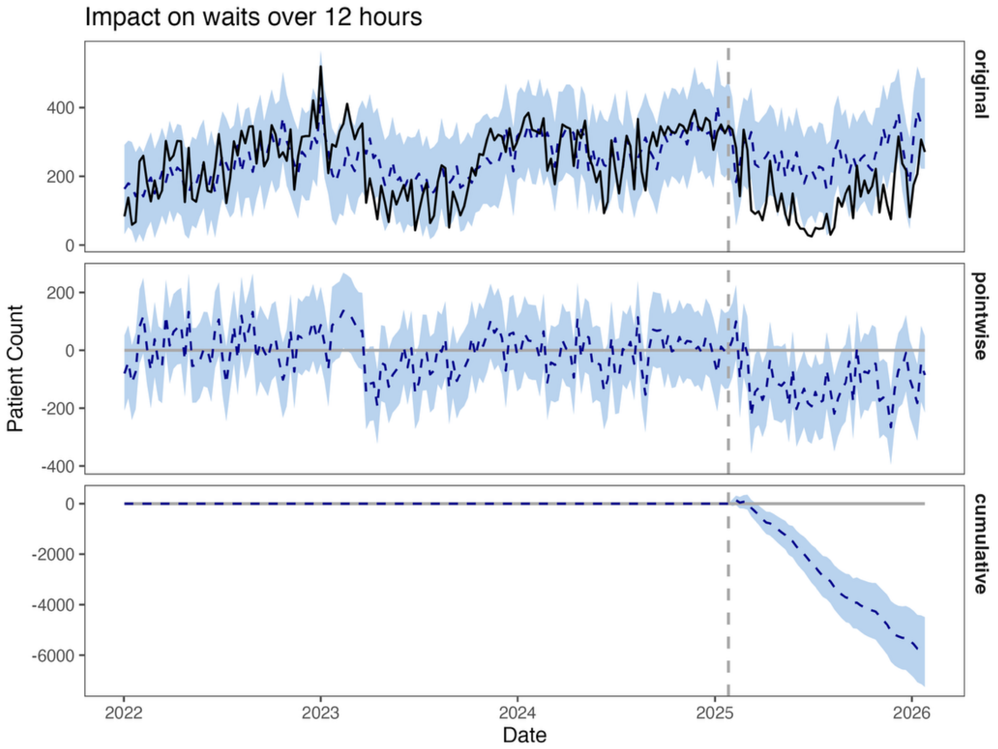
Impact on waits over 12 hours
original
pointwise
cumulative
Patient Count
400
200
0
200
0
-200
-400
0
-2000
-4000
-6000
2022
2023
2024
2025
2026
Date

**4D**

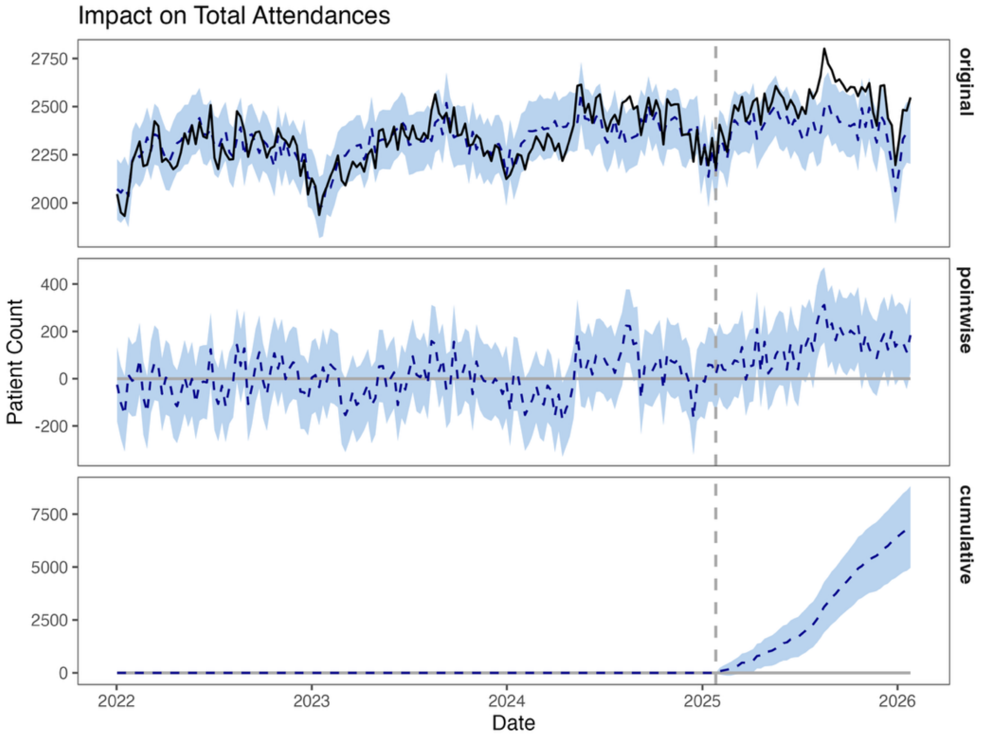
Impact on Total Attendances
original
pointwise
cumulative
Patient Count
2750
2500
2250
2000
400
200
0
-200
7500
5000
2500
0
2022
2023
2024
2025
2026
Date

Supplementary Figure 5. Bayesian changepoint analysis output. Predicted probabilities of structural break.

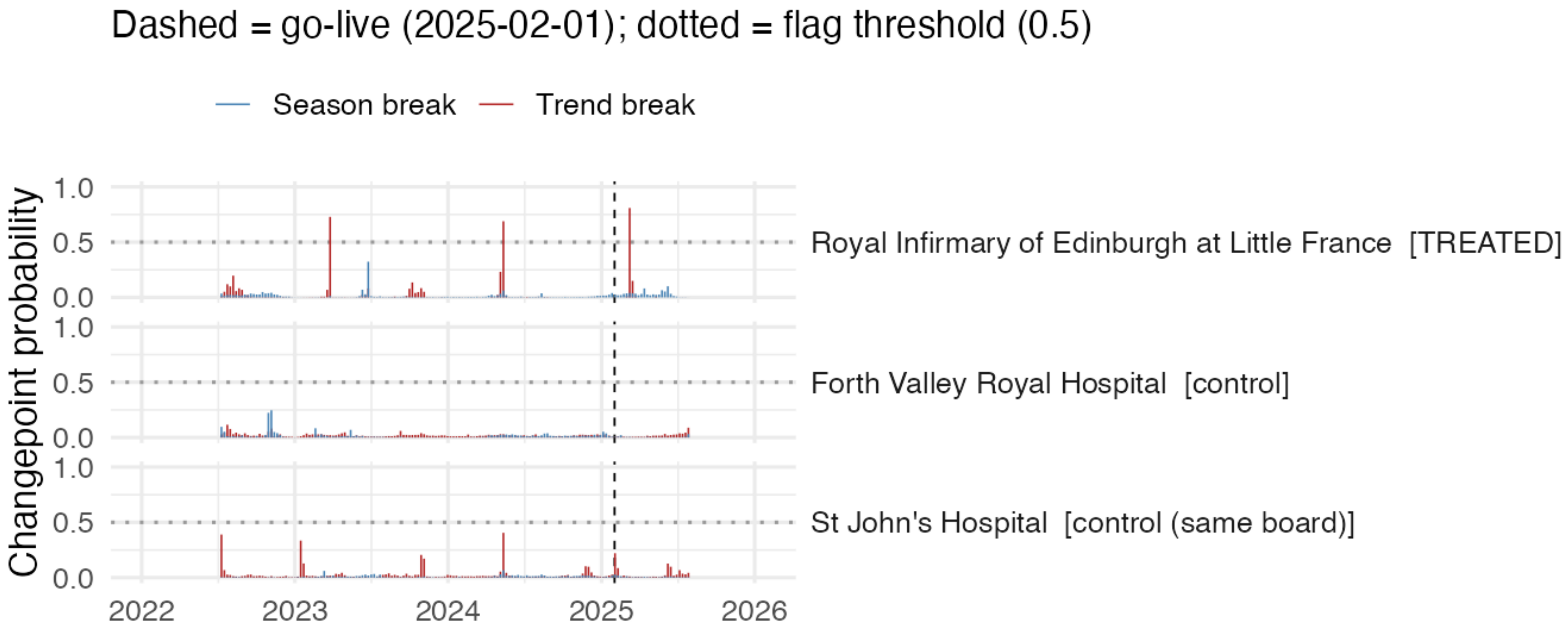

Supplementary Figure 6. Residual autocorrelation and partial autocorrelation function plots for the primary outcome and model.

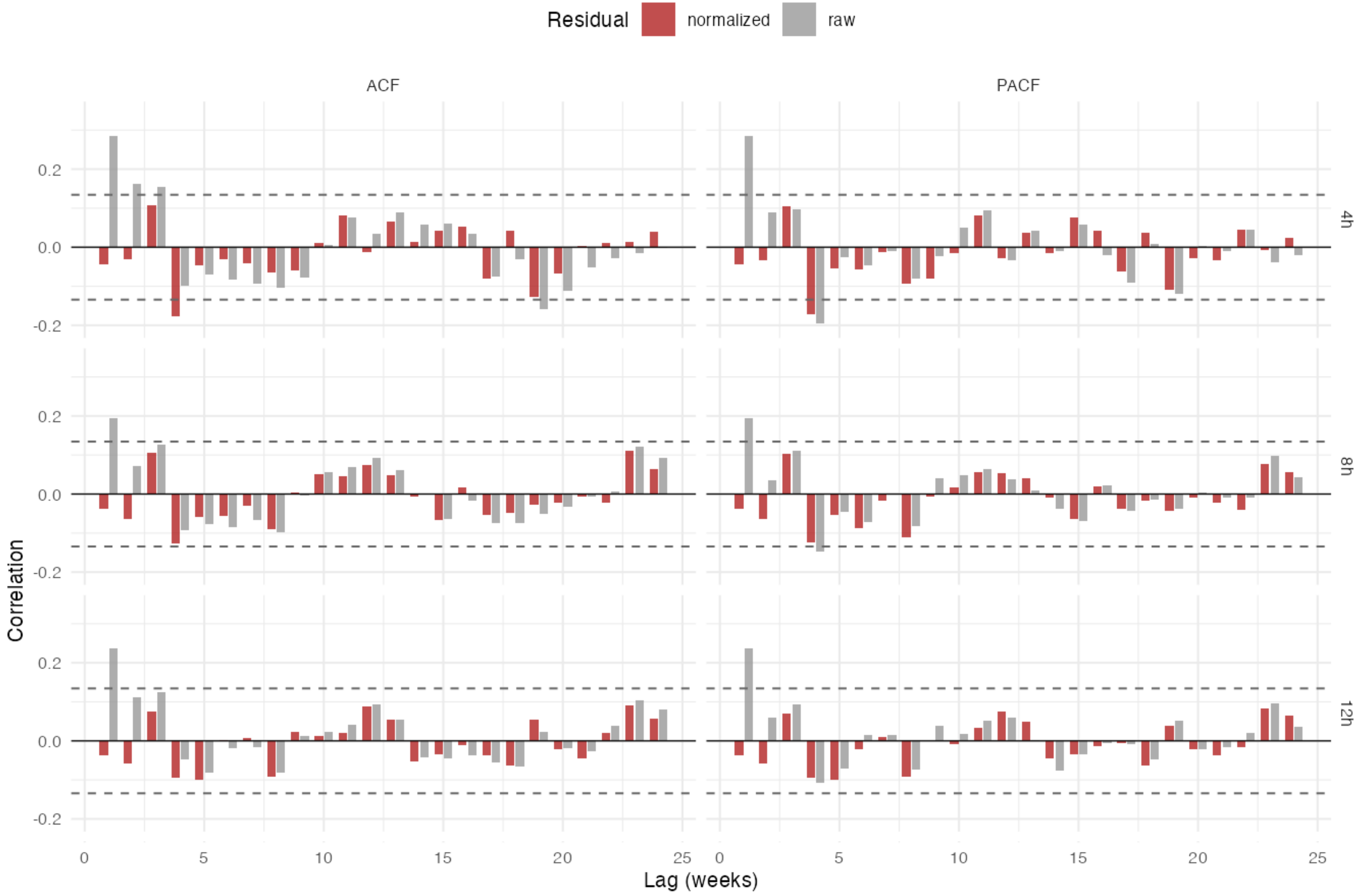
